# An institutional COVID-19 initiative: creation of a biobank and serological data analysis in pre- and post-vaccination cohorts

**DOI:** 10.1101/2024.11.05.24316633

**Authors:** Lorena O Fernandes-Siqueira, Raphael RRA Melo, Luciana S Wermelinger, Fabio CL Almeida, Didier Salmon, Gustavo C Ferreira, Andrea T Da Poian

**Affiliations:** Instituto de Bioquímica Médica Leopoldo de Meis, Universidade Federal do Rio de Janeiro, Rio de Janeiro, RJ, 21941-902, Brazil; Faculdade de Farmácia, Universidade Federal do Rio de Janeiro, Rio de Janeiro, RJ, 21941-902, Brazil

**Keywords:** bank of sera, COVID-19, retrospective observational cohort study, serological surveillance

## Abstract

The COVID-19 pandemic has left a legacy in the management of health emergencies, but sentinel surveillance was relatively underused, despite its significant role in decision-making during epidemics. Here we describe a sentinel SARS-CoV-2 antigens’ serosurveillance carried out on a cohort of 395 individuals at a Brazilian institution, from October 2020 to December 2022. A total of 1,507 serum samples were analyzed for IgG and IgA against SARS-CoV-2 Spike (S) or nucleocapsid (N) proteins, in the pre- and post-vaccination periods. The latter included two doses of CoronaVac (group 1, G1), or ChAdOx-1 or BNT162b2 (group 2, G2), followed by heterologous booster doses. In the pre-vaccination phase, 26.5% of participants showed IgG positivity for S and 13.7% for N. After the vaccines’ first dose, S IgG response was positive in 66.6% or 98% of G1 or G2 participants, respectively, while 100% of the participants showed S IgG positivity after the second dose, and S IgG and IgA after the booster. This initiative enabled the examination of viral transmission beyond hospital environments, which is rarely explored in existing literature, and established protocols for managing future emergencies. In addition, a serum bank and a comprehensive database were established, now available to the scientific community.

## INTRODUCTION

In December 2019, the world faced the biggest health challenge of the last 100 years: the COVID-19 pandemic caused by the new coronavirus SARS-CoV-2 (Tan et al., 2020). Since its onset, 775 million cases and more than 7,000,000 deaths have been reported worldwide (WHO, 2022). Vaccines against COVID-19 were rapidly developed and vaccination began one year after the description of the first cases of the disease (WHO, 2022). Although mass vaccination is the most effective way to control the disease (Fiolet et al., 2022), mitigation policies, such as social distancing, masks usage, hygiene practices, and extensive testing, were enormously important, mainly until high immunization percentages were reached (Güner et al., 2020). Moreover, the need to manage an emerging pandemic that rapidly assumed global proportions required that many measures adopted in other health emergencies, such as influenza outbreaks, were employed (Marcenac et al., 2022).

Sentinel surveillance is a widely used strategy to deal with influenza outbreaks. This strategy is characterized by testing volunteer people across a given community, including those who did not present any disease symptoms, and without necessarily searching for a diagnosis. Its main goal is to discover unseen disease transmission. Although this approach has not been widely used during the COVID-19 pandemic, some sentinel studies carried out in different countries have generated robust epidemiological results that were very useful for decision-making (Vega-Alonso et al., 2023; Cooksey et al., 2021; Miyadahira et al., 2023). In Brazil, for instance, genomic surveillance identified that transition from Delta to the Omicron SARS-CoV-2 variants was faster in healthcare professionals than in other groups of the population (Padilha et al., 2023), while a serosurveillance in low-resource communities in Rio de Janeiro city revealed a seroprevalence much higher than expected in those populations (Brasil et al., 2022; Coelho et al, 2022)

The results obtained from sentinel surveillance become even more significant when there is some difficulty in implementing mitigation strategies, as occurs with workers in essential services who were unable to interrupt their activities during the first months of pandemic, a period characterized by few information and lack of mass diagnosis. Although nowadays the pandemic scenario is different, with more than half of the world’s population vaccinated (WHO, 2022), we must consider that the different initiatives that took place during pandemics also provide valuable learning experience for addressing future emergencies.

In this perspective, our group at the Instituto de Bioquímica Médica Leopoldo de Meis of the Universidade Federal do Rio de Janeiro (IBqM, UFRJ) developed an *in-house* serological test (ELISA) that detects antibodies elicited by SARS-CoV-2 antigens (Fernandes-Siqueira et al., 2022a) to promote serosurveillance of our community. Here, we report a retrospective serological follow-up of a cohort of workers and students from IBqM, UFRJ, spanning the period before the introduction of vaccination in Brazil as well as after the participants undergo different vaccination regimens, namely two doses of CoronaVac, ChAdOx-1 or BNT162b2 as the initial vaccination protocol, followed by heterologous booster doses with ChAdOx-1 or BNT162b2.

## MATERIALS AND METHODS

This work consists of a retrospective observational cohort study, approved by the local ethics committees (CEP approvals HUCFF/UFRJ n° 35.303.120.5.0000.5257). All the participants had access to the informed consent form.

### Study design

The cohort of this study consists of 395 participants, members of IBqM, UFRJ, Brazil, whose serum samples were evaluated to detect antibodies against the spike (S) and nucleocapsid (N) proteins of SARS-CoV-2. The IBqM community was invited by email to participate in the study. All those over 18 years old who accepted the invitation were selected to participate in the study, without exclusion criteria. At the day of the blood collection, they completed an identification form providing information on personal and clinical data. During the 27 months of monitoring (from October 2020 to December 2022), the participants were contacted via a messaging app to schedule blood collection. They had access to their serology results by email. In total, we analyzed 1,507 sera samples from two phases of the pandemic: the pre- and post-vaccination periods.

A total of 242 participants underwent blood collections during the pre-vaccination phase of the study, between October 2020 and April 2021. All these participants had at least one blood sample taken, with a second sample taken for those who tested negative in the first sample, for a total of 313 collections in the whole period. It is important to mention that, although COVID-19 vaccination in Brazil started on January 17, 2021, vaccine application prioritized the population groups more exposed to infection or presenting higher risk of severe disease (healthcare and essential workers, homeless and indigenous people, and elderly and people with chronic health conditions), being extended to the entire population according to age decrease (Santos et al., 2023). As few individuals in our cohort were included in the priority groups, 63% of study participants received the first dose of the vaccine only in April 2021.

In the period between January 2021 to December 2022, 118 participants of the previous group plus 104 new volunteers (total of 222 subjects) participated in the serological monitoring after receiving CoronaVac, ChAdOx-1 or BNT162b2 vaccines against SARS-CoV-2. The general scheme of blood collections in this study phase is presented in Figure 1. All the participants underwent a blood collection 15 to 30 days before receiving the first dose of the vaccine (D1). Post-vaccination collections were divided according to the immunization protocols adopted by the Brazilian Ministry of Health for each vaccine. The blood samples from the CoronaVac group (G1) were collected 20 days after D1, and 20, 60 and 90 days after the second dose (D2). Since the immunization protocol for ChAdOx-1 and BNT162b2 recommended 12 weeks between D1 and D2 (Health Ministry, Brazil, 2024), two blood samples were collected from G2 participants after D1: one between 20 to 45 days and another 80 to 90 days after D1. The third G2 blood sample was collected 15 to 45 days after D2. Both G1 and G2 underwent the following additional collections: 6 months after receiving D1; 20 days and 3 months after receiving the first booster dose (D3); and 20 days after receiving the second booster dose (D4). Therefore, participants vaccinated with CoronaVac underwent a maximum of 8 blood collections and those who received ChAdOx-1 or BNT162b2 underwent a maximum of 7 blood collections (Fig. 1).

**Figure 1.**
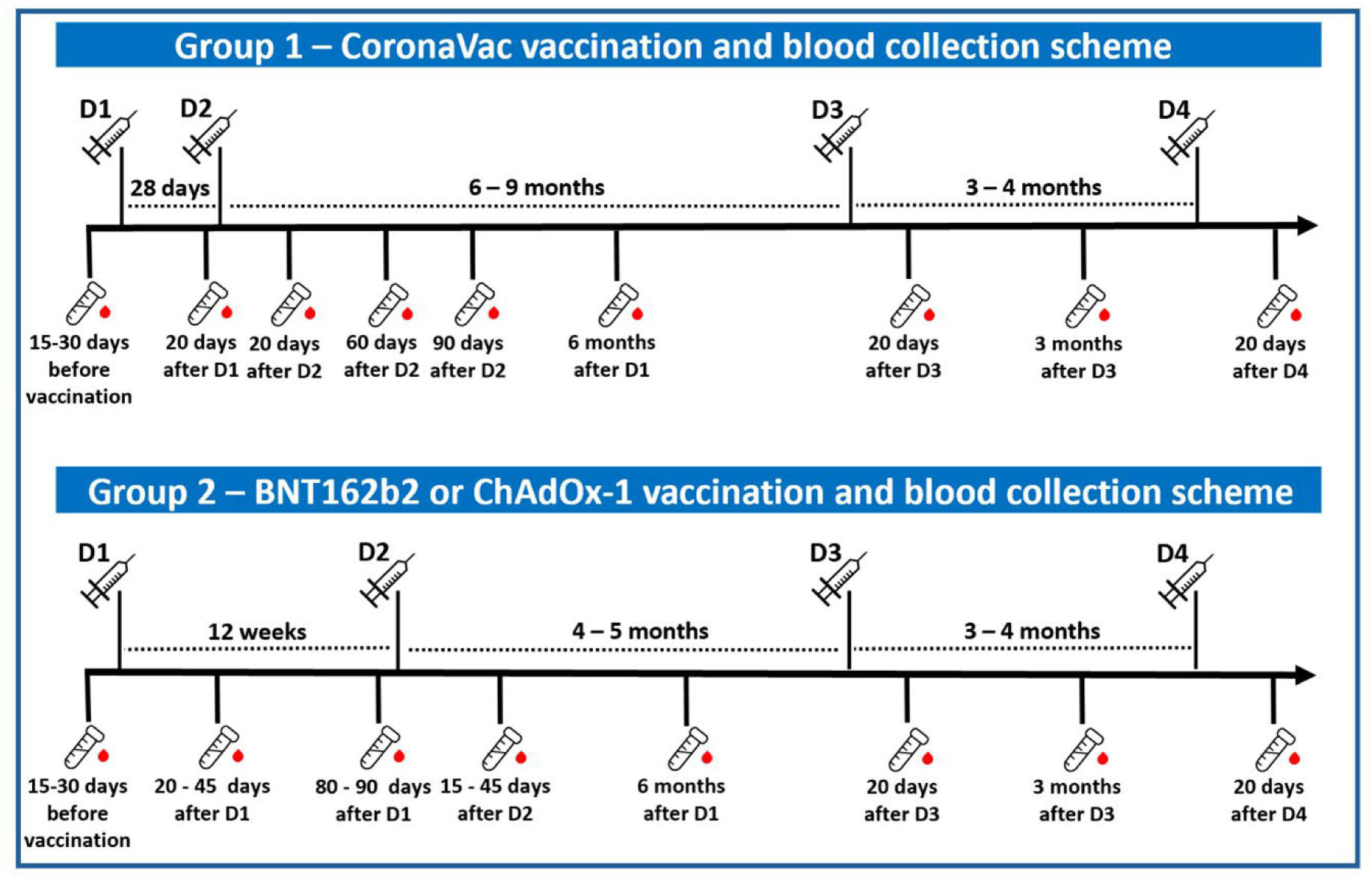
Scheme of blood collections in the post-vaccination period. Schematic representation of blood collections for serological monitoring of participants who received CoronaVac (upper panel), or ChAdOx-1 or BNT162b2 (lower panel).

### Serological analysis (ELISA)

The serological analysis carried out throughout this work were performed using an *in-house* ELISA previously established by our group (Fernandes-Siqueira et al, 2022a). In this assay, we quantified IgG and IgA antibodies against the SARS-CoV-2 trimeric Spike (S) and the N-terminal domain of the nucleocapsid (N) proteins. Briefly, 96 well-plates were previously coated overnight, at 4 °C, with each of the antigens diluted in PBS (50 μL of a 4 μg/mL solution). After 1h blocking with 3% BSA in PBS-T, sera samples, diluted 1:50 in 1% BSA solution in PBS-T, were added to each well and incubated for 2 h, followed by the addition of the detection antibodies (anti-IgG or anti-IgA). The reactivity was quantified spectrophotometrically at 450 nm after the addition of the chromogenic substrate (3,3′,5,5′-tetramethylbenzidine dihydrochloride, TMB). The cut-off for each analysis was calculated as the mean + 3 SD of the absorbance values of 42 pre-pandemic sera.

## RESULTS

The cohort demographic characteristics is presented in Table I. The participants’ median age was 36.5 years old (IQR 27-48), 68% (n=269) of them were women and 32% (n=126) were men. The ethnic-racial self-declaration showed that most individuals consider themselves as “white” (71.6%; n=282), followed by “brown” (20.6%; n=82). Only 6.5% (n=24) self-declared as “black”, 1% (n=4) as “Asian” and 0.25% (1) as “indigenous”. The analysis of participants’ self-reported ABO blood type showed that the predominant blood group in our cohort was type O (40%, n=158), followed by A (35.4%, n=140). B type is less predominant (10.8%, n=21) and AB is the rarest (2.5%, n=10). The blood type distribution in the cohort agrees with that observed in Brazil (World Population Review, 2024). Curiously, 11.1% (n=44) of the participants were either unaware of or chose not to disclose their blood type. A total of 80 participants (20.5%) declared one or more comorbidities, being hypertension the most prevalent (n=42). Of the participants who declared this comorbidity, 47.6% (n=20) were over 60 years, 30.9% (n=13) were between 40 and 59 years, and 21.4% (n=9) were under 40 years (not shown). The second most declared comorbidity was lung diseases (23.7%; n=19), being most of them asthma (n=15) (not shown).

### Pre-vaccination cohort analysis

Using an *in-house* ELISA developed by our group (Fernandes-Siqueira et al., 2022a) we evaluated IgG and IgA responses against SARS-CoV-2 S and N proteins in blood samples collected during the pre-vaccination phase of the study (October 2020 and April 2021, n=242 subjects and 313 samples). Figure 2A shows IgG or IgA reactivity against S or N proteins obtained for each individual, organized in chronological order of blood collections. The results showed a total serum positivity of 26.5% for S IgG, 16% for S IgA, 13.7% for N IgG and 5.1% for N IgA. Data from the identification form filled by the participants revealed that 8,6% had flu-like symptoms and a molecular (PCR) or serological diagnosis for SARS-CoV-2; 34.2% had flu-like symptoms without diagnosis; 55.7% did not have any flu-like symptoms; and 1.2%, although asymptomatic, were PCR-or serology-positive for SARS-CoV-2 (Fig. 2B). We found that 100% of participants who had a positive diagnosis for SARS-CoV-2 infection, 38.8% of the symptomatic participants without a diagnosis, and 11.1% of the asymptomatic participants were positive for S IgG in our test (Fig. 2C). Among the asymptomatic individuals who presented IgG against S, 62.9% also showed IgA reactivity against S, but only 22.2% were positive for N IgG and 11.1% for N IgA, while for symptomatic individuals who tested positive for S IgG, the rate of positivity for S IgA, N IgG and N IgA was 63.4%, 48.7% and 21.9% respectively (Fig. 2D). The double increase in N IgG and IgA could potentially be linked to the onset of symptoms, but further investigation would be required to establish a direct link. Figure 2E shows the main symptoms reported by the symptomatic participants. In agreement with other reports in the literature (Pierron et declared by the participants either previously diagnosed with SARS-CoV-2 infection or showing positive serology in our test. Data provided by the Rio de Janeiro health secretariat (Health Secretariat of The Rio De Janeiro State Government, 2024) showed that between November 2020 and January 2021 there was a pronounced increase in COVID-19 case notifications in the state. This agrees with the seroconversion profile observed for our cohort participants, which reached the highest value in the pre-vaccination period between December 2020 and February 2021 (72 participants) (Fig. 2F). The shift of about one month observed between the two curves in the graph is explained by the fact that the data generated by the Rio de Janeiro’s health secretariat resulted from molecular tests at the onset of symptoms, while participants in our cohort attended blood collection 15 to 30 days after the onset of symptoms.

**Figure 2.**
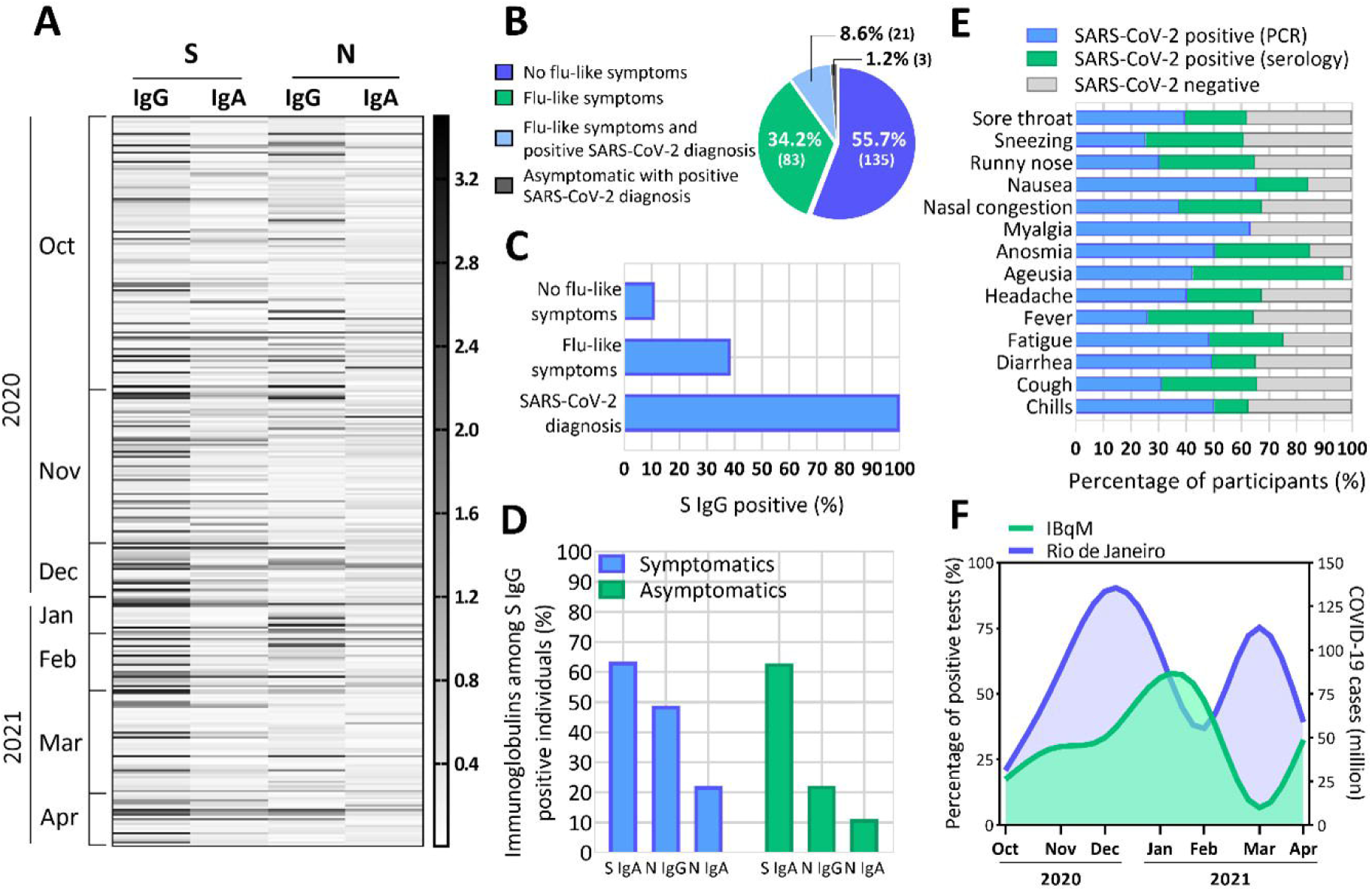
Serological panel and symptomatology of the cohort participants in the pre-vaccination period. (A) Heat map representing sera IgG or IgA reactivity against S or N, detected using an *in-house* ELISA (Fernandes-Siqueira et al., 2022a), in blood samples collected between October 2020 and April 2021. The grey scale used in the heat map represents the optical density (OD) values obtained in the test, being the highest values represented in black, while the values similar to those obtained for the pre-pandemic sera represented in white. The results are organized in chronological order of blood collections. (B) Participants’ grouping according to symptomatology: no flu-like symptoms and no diagnosis (blue); flu-like symptoms without diagnosis (green); flu-like symptoms with previous diagnosis for SARS-CoV-2 (light blue); and asymptomatic with previous diagnosis for SARS-CoV-2 (grey). (C) Participants’ S IgG positivity according with symptomatology and SARS-CoV-2 diagnosis. (D) Positivity for S IgA, N IgG and N IgA among asymptomatic or symptomatic participants who were positive for S IgG. (E) Symptoms declared by the participants showing: molecular diagnosis (PCR+) for SAR-CoV-2 (blue); detection of S IgG by serology (green); or negative result for S IgG (grey). (F) Comparison between the percentage of reactive tests for S IgG in our cohort (green) and the number of confirmed COVID-19 cases in the State of Rio de Janeiro (blue) in the period between October 2020 to April 2021. Data provided by the Health Secretariat of the Rio de Janeiro State Government (2024).

### Post-vaccination cohort analyses

Between January 2021 to December 2022, a total of 222 subjects participated in the serological monitoring after receiving the CoronaVac (n=58) (group 1, G1), or ChAdOx-1 (n=145) or BNT162b2 (n=19) (group 2, G2) vaccines against SARS-CoV-2 (Fig. 3A). The Brazilian vaccination program started with CoronaVac and ChAdOx-1, followed by BNT162b2 and finally Ad26.COV2.S (Health Ministry, Brazil, 2024). In the first months of the vaccination campaign, the majority of the doses administered was of CoronaVac, explaining why most of the individuals of our population received this vaccine from January to March (Fig. 3B). The rapid engagement of Bio-Manguinhos/Fiocruz Immunobiological Technology Institute in the entire production of ChAdOx1 enabled the distribution of this vaccine in Brazil to be largely expanded (Medeiros et al., 2022), explaining the predominance of ChAdOx1 among the vaccines received by our population after March (Fig. 3B). Finally, BNT162b2 was added to the vaccination campaign only in the end of April 2021, resulting in the administration of this vaccine to some participants after May 2021 (Fig. 3B). When inquired about adverse effects they experienced after receiving the vaccines, approximately 70% of the participants who received either the CoronaVac or BNT162b2 vaccines reported no adverse effects following either D1 or D2, while among the individuals who received the ChAdOx-1 vaccine, 59.1% (n=68) declared adverse effects from 24 to 48 hours after the first dose, and 26.9% (n=31) after the second dose (Fig. 3C). Moreover, 20.8% (n=20) of these participants experienced more prolonged adverse effects beyond 48 hours following the first dose of the ChAdOx-1 vaccine. Among the effects, nausea, fatigue, chills, and fever were the most reported (Fig. 3D).

**Figure 3.**
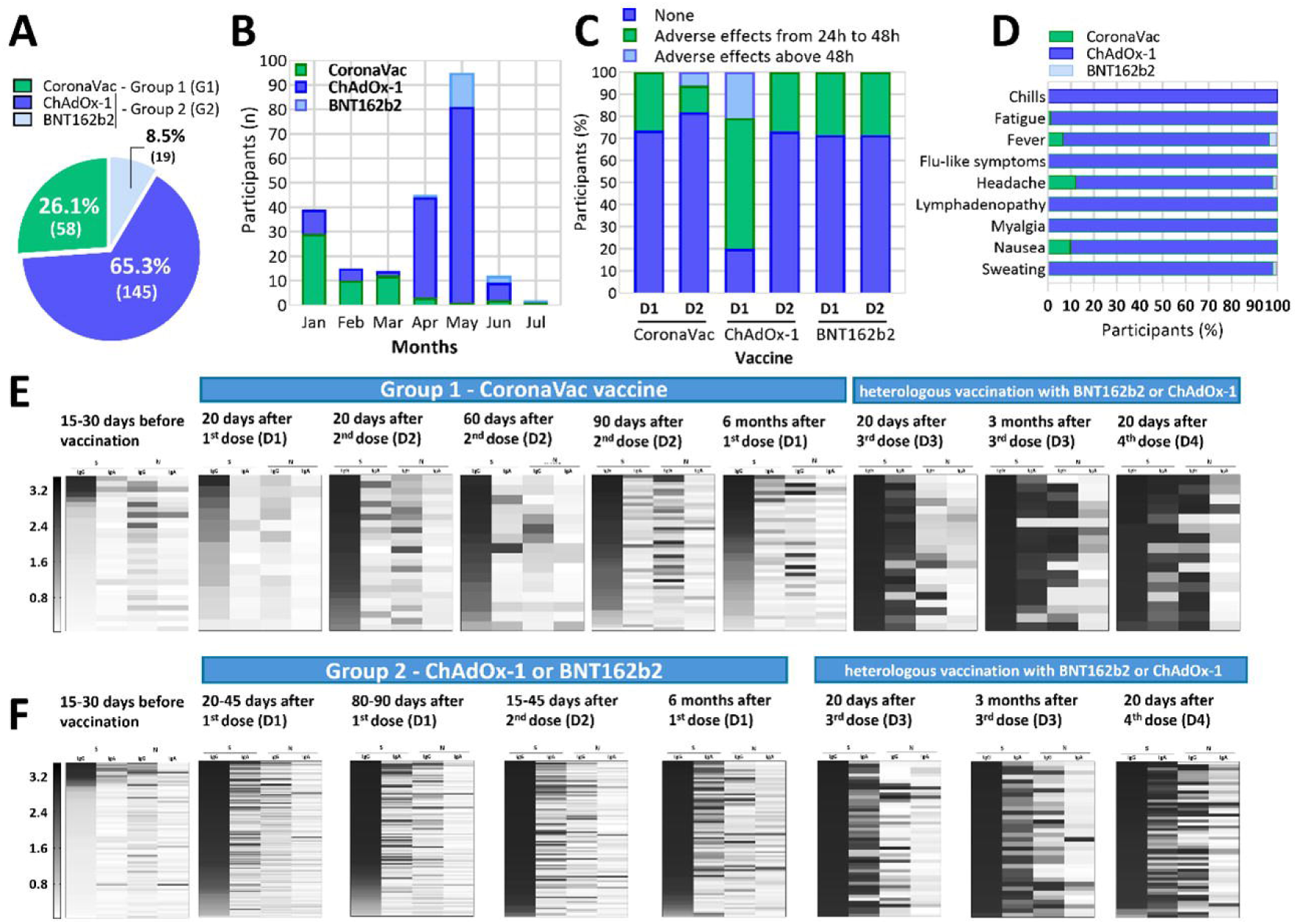
Monitoring of cohort participants in the post-vaccination period. (A) Participants’ grouping according to the vaccine protocol received (first and second dose): CoronaVac (green); ChAdOx-1 (blue); or BNT162b2 (light blue); (B) Vaccine type received as the first dose in each month. (C) Occurrence of adverse effects declared by the participants after receiving the vaccines. (D) Symptoms declared by the participants after receiving the vaccines. (E, F) Heat maps representing sera IgG or IgA reactivity against S or N proteins, detected using an *in-house* ELISA (Fernandes-Siqueira et al., 2022a) in blood samples serially collected (as shown in Fig. 1) between January 2020 and December 2022. The grey scale used in the heat maps represents the optical density (OD) values obtained in the test, being the highest values represented in black, while the values similar to those obtained for the pre-pandemic sera represented in white. The results are organized in ascending order of S IgG OD value (see bars shown on the left). The heat maps are grouped according to the vaccines received: (E) CoronaVac; and (F) ChAdOx-1 or BNT162b2.

Before starting the serological follow-up after vaccination, all participants underwent a blood collection to verify the presence of antibodies against S and N proteins. We observed IgG reactivity to S in 20% of G1 participants (Fig. 3E) and 25% of G2 participants (Fig. 3F), indicating that these individuals underwent previous infection with SARS-CoV-2. After the first dose of the vaccines (D1), S IgG has been detected in 66.6% of the G1 and 98% of G2 participants. On the other hand, S IgA was detected in only 4% or 6.9% of the G1 or G2 participants, respectively. After the second dose of the vaccines (D2), the percentage of S IgG detection increased to 100% in both groups, while S IgA appeared in 66% of the G1 and 96.9% of the G2 participants. Six months after receiving D1, detection of S IgG dropped to 87.5% in G1 participants, while IgG reactivity to S was maintained in 100% in G2 individuals (Figs. 3E-F).

Previous studies have shown that immunization with the three vaccines used in this work, including CoronaVac, which uses the whole inactivated virus, does not elicit a significant serological response against the N protein (Bochnia-Bueno et al., 2022; Huergo et al., 2022). Therefore, the detection of N IgG in the population analyzed in the vaccination cohort may be an indicative of new cases of COVID-19 during the vaccination period. Before vaccination, 17.2% of the G1 participants and 11.6% of the G2 participants presented N IgG, while six months later, the percentage of N IgG positivity increased to 20% and 15.2% in G1 and G2, respectively. This suggests a low incidence of infection among vaccinated individuals, as the increase in N IgG positivity over time may be attributed to new cases rather than vaccine induced responses.

All participants who participated in post-vaccination monitoring in this study received the heterologous first booster dose (D3), following the immunization protocol adopted by the Brazilian Ministry of Health. This protocol involved individuals vaccinated with CoronaVac received either ChAdOx-1 or BNT162b2 vaccines, those vaccinated with ChAdOx-1 receiving BNT162b2, and those vaccinated with BNT162b2 receiving ChAdOx-1. The booster dose was very effective in inducing the humoral immune response, with 100% of participants in both groups testing positive for S IgG and IgA. This high percentage was maintained up to three months after D3 and after receiving the second booster dose (D4). Although these high levels of antibodies against SARS-CoV-2 attest that the booster dose promoted a very effective immunization, increased rates of N IgG were observed after D3 and D4 (66.6% and 76.4% in G1, and 51.4% and 62.9% in G2), suggesting a rise in COVID-19 cases during this period. However, no participant reported severe symptoms (data not shown).

## DISCUSSION

Viral elimination and mitigation policies during the first year of COVID-19 pandemic were the most important strategies for controlling the disease (Oliu-Barton et al., 2021). Although the presence of antibodies against SARS-CoV-2 in the pre-vaccination period did not assure protection to justify the relaxation of quarantine or social distancing measures (West et al., 2021), the rapid implementation of serological tests is an important tool for surveillance of viral spread and for the management of health emergencies (Novello et al., 2021). Here, we followed the serological response against SARS-CoV-2 antigens in a cohort of 395 healthy individuals over a 27-months period, before and after the vaccination. Although our cohort represents almost 80% of the entire IBqM social body, the small number of participants would represent a limitation for some data interpretation. Nevertheless, here we monitored seroprevalence outside hospital environments, which is unusually reported in the literature specially before COVID-19 vaccines became available, representing important sentinel-like surveillance that tracks viral spread even in asymptomatic individuals.

The results obtained during the pre-vaccination period of our study indicated a low incidence of individuals with positive serology (26.1%) in the first year of the pandemic, as also reported for a similar cohort (Brasil et al., 2022; Coelho et al., 2022; Henriques-Santos et al., 2022) Nevertheless, we identified 11% of positive cases among the asymptomatic individuals, which supported the recommendation of molecular testing and social distancing independently of the presence of symptoms. Indeed, the increase in seroconversion between December 2020 and February 2021 seemed to be related to the relaxation of protective measures during festive periods (Geenen et al., 2023), as well as the emergence of the first variant of concern, P1 (Carvalho et al. 2022). Additionally, the correlation between this observation and the increase in COVID-19 cases in the State of Rio de Janeiro reinforces the usefulness of serological tests to alert the growing number of infected people. It is also important to mention that, during the first year of the pandemic, the number of serological tests carried out worldwide, and mainly in Brazil, was low due to their high cost and limited availability (West et al., 2021) In this context, the development of a lower-cost *in-house* ELISA (Fernandes-Siqueira et al., 2022a) by our group was crucial for conducting the tests during that period.

Safety and efficiency of the vaccination regimens using CoronaVac, ChAdOx-1, and BNT162b2, as well as the complementary booster doses, have been demonstrated in studies carried out in Brazil and other countries (Bochnia-Bueno et al., 2022; De Sousa et al., 2022; Shrotri et al., 2022; Paula et al., 2022). ChAdOx-1 and BNT162b2 promoted a stronger immune response when compared to CoronaVac (Romero-Ibarguengoitia et al. 2022; Lin-Wang et al., 2022; Fernandes-Siqueira et al., 2022b), although our data showed that two doses of all three vaccines provided a good humoral response in our cohort. After D3 and D4, anti-S antibody levels remained quite high across the entire cohort, regardless of the vaccination regimen, consistent with findings from other studies (Da Penha Gomes Gouvea et al., 2023; Yechezkel et al., 2023). Remarkably, the considerable increase in anti-N antibodies during this period possibly reflects the emergence of the highly transmissible Omicron variant (Jalali et al., 2022; Murari et al., 2022).

In conclusion, this study supports that monitoring seroconversion in a cohort composed of healthy individuals during a pandemic situation, before and after the implementation of different vaccination regimens, is a strategy that can be applied in the surveillance of immune coverage in new situations. This strategy can be applied to both small centers and larger populations, considering the regional differences and government measures (Fawole et al., 2023; Muttamba et al., 2022). Our work also generated a biobank with over 1,500 sera samples, resulting from up to 8 blood collections from the same individual during different pandemic phases. Furthermore, this biobank, as well as its respective demographic and serological data, could be useful for the scientific community in future health emergencies.

## Data Availability

All data produced in the present study are available upon reasonable request to the authors.

## ACKNOWLEDGEMENTS

We would like to thank Lavinia Reif Correa de Oliveira and Lilian Cristine de Ferreira (UFRJ, Brazil) for helping with sample organization; Dr. Marcos Fleury and all the members of Laboratório de Análises Clínicas, Faculdade de Farmácia (LACFar, UFRJ, Brazil) for performing the blood collections and sample preparation; our colleagues from Instituto de Bioquímica Médica Leopoldo de Meis (IBqM, UFRJ, Brazil), for financial support and encouragement; all the members of Laboratório de Bioquímica de Vírus for providing recombinant N protein; and Dr. Leda Castilho research group (COPPE, UFRJ, Brazil) for kindly providing recombinant S protein.

This work was supported by Fundação Carlos Chagas Filho de Amparo à Pesquisa do Estado do Rio de Janeiro (FAPERJ), Brazil [grant numbers E-26/211.128/2021, E-26/201.173/2021]; and Conselho Nacional de Desenvolvimento Científico e Tecnológico (CNPq), Brazil [grant number 312650/2021-3].

## AUTHORS’ CONTRIBUTIONS

LOFS: conceptualisation, investigation, methodology, data curation, formal analysis, writing - original draft, review & editing. RRAM: methodology, data curation, formal analysis. LSW: conceptualisation, investigation, methodology, writing — original draft, review & editing. FCLA: conceptualization, resources. DS: conceptualization; writing: review & editing. GCF: conceptualisation, investigation, writing - review & editing. ATDP: conceptualization, investigation, writing: review and editing, resources, funding acquisition, supervision, project administration. All authors read and approved the final manuscript.

